# Small Airways Disease is a Post-Acute Sequelae of SARS-CoV-2 Infection

**DOI:** 10.1101/2021.05.27.21257944

**Authors:** Josalyn L. Cho, Raul Villacreses, Prashant Nagpal, Junfeng Guo, Alejandro A. Pezzulo, Andrew L. Thurman, Nabeel Y. Hamzeh, Robert J. Blount, Spyridon Fortis, Eric A. Hoffman, Joseph Zabner, Alejandro P. Comellas

## Abstract

**Background:** The sequelae of SARS-CoV-2 infection on pulmonary structure and function remain incompletely characterized.

**Methods:** Adults with confirmed COVID-19 who remained symptomatic more than thirty days following diagnosis were enrolled and classified as ambulatory, hospitalized or requiring the intensive care unit (ICU) based on the highest level of care received during acute infection. Symptoms, pulmonary function tests and chest computed tomography (CT) findings were compared across groups and to healthy controls. CT images were quantitatively analyzed using supervised machine-learning to measure regional ground glass opacities (GGO) and image-matching to measure regional air trapping. Comparisons were performed using univariate analyses and multivariate linear regression.

**Results:** Of the 100 patients enrolled, 67 were in the ambulatory group. All groups commonly reported cough and dyspnea. Pulmonary function testing revealed restrictive physiology in the hospitalized and ICU groups but was normal in the ambulatory group. Among hospitalized and ICU patients, the mean percent of total lung classified as GGO was 13.2% and 28.7%, respectively, and was higher than in ambulatory patients (3.7%, P<0.001). The mean percentage of total lung affected by air trapping was 25.4%, 34.5% and 27.2% in the ambulatory, hospitalized and ICU groups and 7.3% in healthy controls (P<0.001). Air trapping measured by quantitative CT correlated with the residual volume to total lung capacity ratio (RV/TLC; ρ=0.6, P<0.001).

**Conclusions:** Air trapping is present in patients with post-acute sequelae of COVID-19 and is independent of initial infection severity, suggesting obstruction at the level of the small airways. The long-term consequences are not known.

## Introduction

Severe acute respiratory syndrome coronavirus 2 (SARS-CoV-2), the virus that causes COVID-19, primarily infects the respiratory tract and causes a wide range of disease severity, including respiratory failure due to the acute respiratory distress syndrome (ARDS)^1^. Survivors of severe COVID-19 have pulmonary function abnormalities that persist for weeks to months after resolution of the acute illness^2,3^. Additionally, interstitial changes are frequently observed on lung imaging following severe infection. These findings are consistent with literature on the long-term respiratory sequelae of ARDS^4,5^ and observations from previous severe coronavirus outbreaks^6,7^.

Early reports of post-acute sequelae of COVID-19 (PASC), or “long COVID”, suggest that respiratory symptoms, including cough and dyspnea, are common among COVID-19 survivors and that they occur in a significant proportion of patients who experienced mild infection not requiring hospitalization^8,9^. The long-term effects of SARS-CoV-2 infection in these patients are poorly understood, but the potential impact on our healthcare systems is enormous given the millions of infections worldwide, most of whom had mild disease. The primary aim of this study was to characterize the effects of SARS-CoV-2 on pulmonary structure and function in the post-acute period. Specifically, we sought to determine whether respiratory symptoms, pulmonary function or radiographic abnormalities varied based on the severity of acute infection.

## Methods

### Patients

In response to the COVID-19 pandemic, the University of Iowa established a post-acute COVID-19 outpatient clinic for patients with confirmed SARS-CoV-2 infection who remained symptomatic more than thirty days following diagnosis. Eligible patients were adults aged 18 or older referred to the post-COVID-19 outpatient clinic who provided written informed consent for participation in a research registry and collection of their clinical data. Confirmed COVID-19 was defined as a positive result on either a rapid antigen test or a reverse-transcriptase-polymerase-chain-reaction (RT-PCR) test from a nasopharyngeal or oropharyngeal swab or a positive SARS-CoV-2 antibody test. The study was approved by the institutional review board of the University of Iowa. The authors analyzed only de-identified data.

### Study Design, Definitions and Assessments

The period of acute COVID-19 infection was defined as the twenty-one days following diagnosis. Patients were classified as ambulatory, hospitalized but not requiring the intensive care unit (hereafter referred to as hospitalized) and requiring the intensive care unit (ICU) based on the highest level of care received during acute COVID-19 infection. Demographic, clinical and laboratory data from the period of acute COVID-19 was collected from electronic medical records. During the clinic visit, patients underwent a comprehensive history and physical examination. Patients were asked to retrospectively recount their symptoms during the acute phase of infection and detail whether these symptoms persisted using a structured questionnaire (Supplementary Appendix). The modified Medical Research Council (mMRC) dyspnea scale was used to quantify breathlessness, where 0 indicates dyspnea only with strenuous exercise and 4 indicates dyspnea when dressing^10,11^. All patients underwent laboratory testing, pulmonary function testing and chest imaging on the day of their clinic visit as part of the clinical protocol. Pulmonary function testing and chest CT data was compared to a cohort of healthy non-smoking control subjects (Supplementary Appendix), who were matched to post-COVID-19 patients based on age, sex and ethnicity (**Table S1**). We pre-specified that analysis would be performed after the first 100 patients were consented.

### Chest Computed Tomography Acquisition and Image Analysis

Patients and healthy subjects underwent non-contrast chest CT with an inspiratory scan coached to total lung capacity (TLC) and an expiratory scan coached to residual volume (RV). The scanning protocol for all subjects was standardized as previously described and adhering to the principles outlined by the Quantitative Imaging Biomarker Alliance^12,13^. Qualitative CT image analysis was performed by an experienced, blinded radiologist (P.N.). CT images were also quantitatively analyzed using texture analysis (Adapted Multiple Feature Method; AMFM) and Disease Probability Measure (DPM, VIDA Diagnostics). These machine-learning- and image-matching-based approaches are detailed in the Supplementary Appendix^12,14–19^. Briefly, the AMFM-based texture analysis quantifies ground glass opacities (GGO) as a percent of total lung volume at TLC by utilizing grayscale patterns within CT images. DPM quantifies the voxel-to-voxel difference in Hounsfield Units (HU) between matched inspiratory and expiratory images to estimate the probability of air trapping such that the probability is inversely proportional to the relative differences in HU^19^. When quantified using DPM analysis, air trapping reflects functional small airways disease (fSAD)^19–21^.

### Statistical Analysis

Data are summarized using descriptive statistics. Continuous variables are reported using medians and interquartile ranges or means and standard error measurements. Categorical variables are reported as counts and percentages. No imputation was made for missing data. Analyses were pre-specified to determine whether there were differences across the ambulatory, hospitalized and ICU groups and between the ambulatory group and healthy controls. Univariate comparisons were made using the Chi-squared test, one-way ANOVA or the Mann-Whitney test; the Kruskal-Wallis test was used to correct for multiple comparisons across groups. Multivariable linear regression was used to adjust for age and body mass index (BMI) and the least square means were calculated; correction for multiple comparisons across groups was performed using Tukey’s test. Spearman’s correlation coefficients were used to assess the strength of association between pairs of predefined variables. In all cases, differences were considered statically significant when *P* was less than 0.05 based on a two-sided test. Statistical analyses were performed using GraphPad Prism or R statistical software (http://www.r-project.org).

## Results

### Demographic and Clinical Characteristics of the Participants at Baseline

Between June 2020 and January 2021, 110 patients were evaluated in the post-acute COVID-19 clinic and were eligible for enrollment. 101 individuals (91.8%) consented to participate. One patient was excluded because the diagnosis of COVID-19 could not be confirmed. Thus, 100 individuals were included in the analysis.

The demographic and clinical characteristics of the participants at baseline are shown in **Table 1**. Most patients were treated in the ambulatory setting (67%) during the acute COVID-19 period. The median age of the patients was 48 years (IQR, 36.3 to 60.5 years) and 66% were female. Patients who were hospitalized or in the ICU were significantly older than ambulatory patients (P<0.001 and P<0.01, respectively). At least one co-existing illness was present in 76 patients. Obesity (59%) and hypertension (27%) were the most common co-existing illnesses. Critically ill patients were more likely to have chronic kidney disease, chronic obstructive pulmonary disease, type 2 diabetes mellitus and hypertension compared to ambulatory patients. The most common co-existing pulmonary disorder was asthma (26%), followed by chronic obstructive pulmonary disease (6%) and interstitial lung disease (4%). The majority of patients were never smokers (75%). Smoking was more common among hospitalized or ICU patients than ambulatory patients. Among former and current smokers, median pack-years were 8 (IQR, 4.8-25.5). Median time to follow-up in the post-COVID ambulatory clinic was 74.5 days (IQR, 45.8-118).

**Table 1:** Demographic and clinical characteristics of the patients at baseline.

|  |  |  |  |  |
| --- | --- | --- | --- | --- |
| **Characteristic** | **All Patients**<br>**(N=100)** | **Ambulatory**<br>**(N=67)** | **Hospitalized**<br>**(N=17)** | **ICU**<br>**(N=16)** |
| Age, median (IQR) - yr | 48 (36.3-60.5) | 44 (29-53) | 64 (52.5-71.5)<sup>***</sup> | 60 (45-68)<sup>**</sup> |
| Sex - no. (%) |  |  |  |  |
| Male | 34 (34%) | 19 (28.4%) | 6 (35.3%) | 9 (56.3%) |
| Female | 66 (66%) | 48 (71.6%) | 11 (64.7%) | 7 (43.8%) |
| Race and ethnic group - no. (%)<sup>#</sup> |  |  |  |  |
| White, non-Hispanic | 85 (85%) | 59 (88.1%) | 14 (82.4%) | 12 (92.3%) |
| Black, non-Hispanic | 5 (5%) | 3 (4.5%) | 1 (5.9%) | 1 (7.7%) |
| Hispanic or Latino | 10 (10%) | 5 (7.5%) | 2 (11.8%) | 3 (18.8%) |
| Body-mass index, median (IQR)<sup>§</sup> | 32<br>(26.6-38.1) | 31<br>(23.7-36.6) | 36<br>(30.1-48.2)<sup>*</sup> | 36<br>(26.7-38) |
| Coexisting disorder - no. (%) |  |  |  |  |
| Asthma | 26 (26%) | 18 (26.9%) | 4 (23.5%) | 4 (25%) |
| Cancer | 9 (9%) | 5 (7.5%) | 3 (17.6%) | 1 (6.3%) |
| Chronic kidney disease | 4 (4%) | 0 (0%) | 1 (5.9%) | 3 (18.8%)<sup>**</sup> |
| Chronic obstructive pulmonary disease | 6 (6%) | 1 (1.5%) | 1 (5.9%) | 4 (25%)<sup>**</sup> |
| Coronary artery disease | 8 (8%) | 4 (6%) | 2 (11.8%) | 2 (12.5%) |
| Diabetes type I | 2 (2%) | 1 (1.5%) | 1 (5.9%) | 0 (0%) |
| Diabetes type II | 12 (12%) | 3 (4.5%) | 3 (17.6%) | 6<br>(37.5%)<sup>***</sup> |
| Heart failure | 1 (1%) | 0 (0%) | 1 (5.9%) | 0 (0%) |
| Hypertension | 27 (27%) | 10 (14.9%) | 8 (47.1%)* | 9 (56.3%)** |
| Immunocompromise or HIV | 8 (8%) | 3 (4.5%) | 3 (17.6%) | 2 (12.5%) |
| Interstitial lung disease | 4 (4%) | 2 (3%) | 2 (11.8%) | 0 (0%) |
| Obesity | 59 (59%) | 36 (53.7%) | 13 (76.5%) | 10 (62.5%) |
| Thromboembolic disease | 3 (3%) | 0 (0%) | 2 (11.8%) | 1 (6.3%) |
| Tobacco use - no. (%) |  |  |  |  |
| Never | 75 (75%) | 57 (85.1%) | 8 (47.1%***) | 10 (62.5%)* |
| Former | 23 (23%) | 8 (11.9%) | 9 (52.9%) | 6 (37.5%) |
| Current | 2 (2%) | 2 (3.0%) | 0 (0%) | 0 (0%) |
| Pack-yr, median (IQR)<sup>†</sup> | 8 (4.8-25.5) | 12 (5-27.5) | 5.5 (3.1-20.5) | 10 (2.5-40) |
| Time to follow-up - days, median (IQR)<sup>‡</sup> | 74.5 (45.8-118) | 68 (42-126) | 109 (70.5-204) | 59.5 (50.3-97.8) |

*Abbreviations:* HIV, human immunodeficiency virus; IQR, interquartile range
<sup>^</sup>Percentages may not total 100 because of rounding.
<sup>#</sup>Race and ethnic group were reported by the patient.
<sup>§</sup>The body-mass index (BMI) is the weight in kilograms divided by the square of the height in meters.
<sup>†</sup>Pack-years are only calculated for former and current smokers. One pack-year is defined as smoking one package of cigarettes per day on average for one year.
<sup>‡</sup>Time to follow-up is defined as time from laboratory confirmed diagnosis to the post-COVID-19 clinic visit.
\*P<0.05, \*\*P<0.01, and \*\*\*P<0.001 compared to ambulatory patients.

### Clinical Characteristics of Acute and Post-Acute COVID-19

The most common symptoms during acute COVID-19 infection were fatigue (83.7%), dyspnea (82.3%), and cough (71.4%) (**Figure 1, Table S2**). The median length of admission was 4 days (IQR, 2.5-7) for the hospitalized group and 18.5 days (IQR, 10.8-42.3) for the ICU group (P<0.001). None of the ambulatory patients required supplemental oxygen during acute illness. Ten (58.8%) of the hospitalized patients and all of the critically ill patients required supplemental oxygen. Among hospitalized patients requiring supplemental oxygen, the median maximal requirement was 2 liters per minute (IQR, 0-2). Of the ICU patients, 14 (87.5%) required high-flow oxygen, 11 (68.8%) were mechanically ventilated and 3 (18.8%) required extracorporeal membrane oxygenation (ECMO). The median maximal fraction of inspired oxygen for ICU patients was 1.0 (IQR, 0.65-1.0). Additional details of the ICU-level therapies received by critically ill patients and laboratory values during acute COVID-19 are provided in **Table S2**.

**Figure 1:**
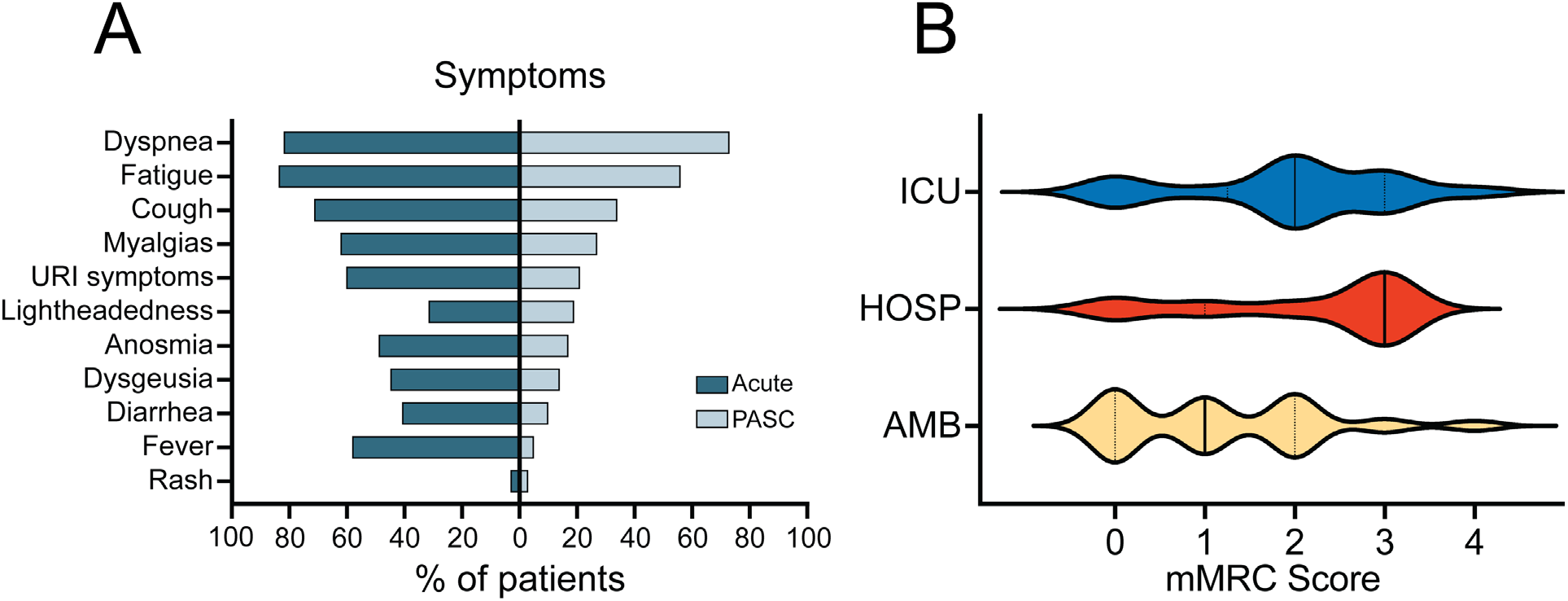
Symptoms during post-acute COVID-19. (Panel A) Frequency of symptoms reported during acute COVID-19 (left; dark blue bars) and post-acute COVID-19 (right; light blue bars). (Panel B) Severity of dyspnea by group quantified using the modified Medical Research Council (mMRC) scale. *Abbreviations*: AMB, ambulatory; HOSP, hospitalized; ICU, intensive care unit

The most common treatments for COVID-19 were corticosteroids (36%), antibiotics (18%) and remdesivir (16%) (**Table S2**). Hospitalized and ICU patients were more likely than ambulatory patients to receive treatment with corticosteroids (64.7%, 87.5% and 16.4%, respectively; P<0.001 for both comparisons), remdesivir (29.4%, 68.8% and 0%; P<0.001 for both comparisons) and convalescent plasma (17.6%, 56.3% and 1.5%; P<0.05 and P<0.001, respectively). Two patients each in the ambulatory (3%) and hospitalized (11.8%) groups and one patient (6.3%) in the ICU group experienced venous thromboembolism as a complication of acute SARS-CoV-2 infection. Additional complications, including the need for renal replacement therapy (12.5%), pneumothorax (6.3%) and the need for tracheostomy (18.8%) were observed in the ICU group.

Similar to acute infection, the most commonly reported persistent symptoms at follow-up were dyspnea (73%), fatigue (56%) and cough (34%) (**Figure 1, Table S3**). At follow-up, two patients in the hospitalized group (11.8%) and 9 patients in the ICU group (56.3%) had an ongoing requirement for supplemental oxygen that was not present prior to SARS-CoV-2 infection. Additional clinical data for post-acute COVID-19 is provided in **Table S3**.

### Outcomes

The median score on the mMRC dyspnea scale at follow-up was 2 (IQR, 0-2) (**Figure 1, Table S4**). Compared to the ambulatory group (38.8%), a higher proportion of patients in the hospitalized (70.6%) and ICU (75%) groups had an mMRC score greater than or equal to 2 (P<0.01). The median percent predicted pre-bronchodilator forced vital capacity (FVC) and forced expiratory volume in one second (FEV1) in post-acute COVID-19 patients were 95% (IQR 81-106) and 93% (IQR 78-104), respectively (**Figure 2**). Compared to the ambulatory group, the hospitalized and ICU groups had a lower FVC (P<0.01 and P<0.001, respectively) and FEV1 (P<0.05 and P<0.001, respectively). The FVC and FEV1 in ambulatory patients were not different compared to healthy controls. The median pre-bronchodilator FEV1/FVC was 0.8 (IQR, 0.76-0.84), and there were no differences across groups. No clinically significant response to bronchodilator was observed for FVC or FEV1 in any of the PASC groups. Median percent predicted total lung capacity (TLC) was 96% (IQR, 83.5-109) and median residual volume was 82% (IQR, 64.3-98.8) in PASC patients. Compared to ambulatory patients, hospitalized and critically ill patients had lower TLC (P<0.01 and P<0.001, respectively). Critically ill patients also had lower RV compared to ambulatory patients (P<0.01). TLC and RV were similar between the ambulatory group and healthy controls. The median percent predicted diffusing capacity for carbon monoxide (DLCO) was 96% (IQR, 79-111.5) in PASC patients. Hospitalized and critically ill patients had a significantly lower DLCO compared to ambulatory patients (P<0.001 for both comparisons). The DLCO was significantly higher in ambulatory patients compared to healthy controls (P<0.001). Similar differences in spirometry, lung volumes and DLCO were found in our age- and BMI-adjusted multivariate analysis (**Figure S1**).

**Figure 2:**
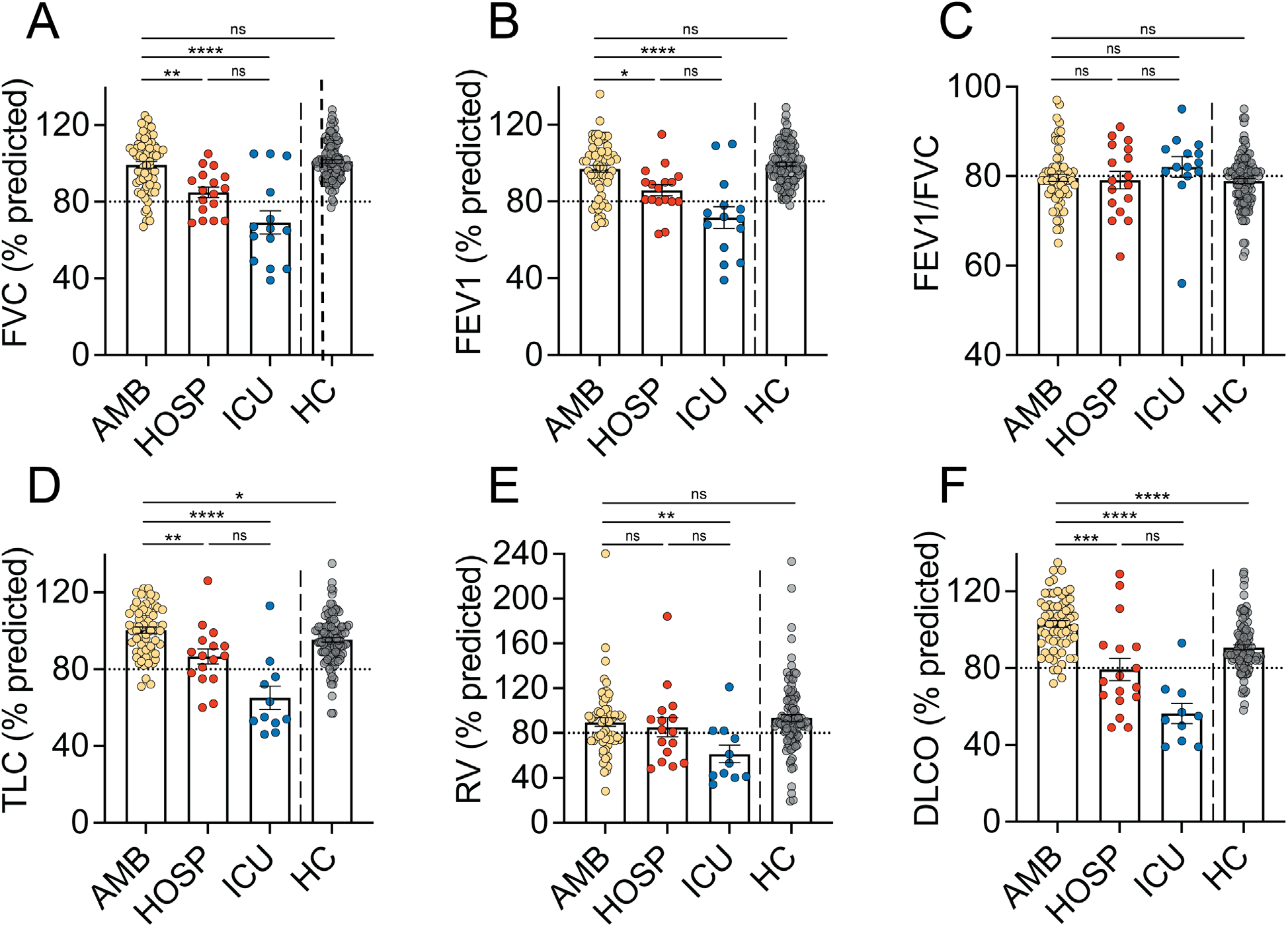
Pulmonary function testing in post-acute COVID-19. Shown are the pulmonary function testing data by group. (Panel A) Percent predicted forced vital capacity (FVC). (Panel B) Percent predicted forced expiratory volume in 1 second (FEV1). (Panel C) FEV1/FVC. (Panel D) Percent predicted total lung capacity (TLC). (Panel E) Percent predicted residual volume (RV). (Panel F) Percent predicted diffusion capacity for carbon monoxide. Data are displayed as mean with standard error measurement (SEM). Horizontal dashed lines indicate the lower limit of normal. *P<0.05, **P<0.01 and ***P<0.001. *Abbreviations*: AMB, ambulatory; HOSP, hospitalized; ICU, intensive care unit; HC, healthy controls

Images from inspiratory and expiratory chest CT were available for analysis for 91 patients (91%). The most common abnormalities identified by qualitative analysis were air trapping (58%), GGO (51%), and pulmonary nodules (35%) (**Table 2**). Representative chest CT images demonstrating GGO, air trapping and architectural changes are shown in **Figure S2**. Hospitalized and critically ill patients were more likely to have bronchiectasis and architectural distortion, honeycombing or scar compared to ambulatory patients. To more fully assess GGO and air trapping, quantitative analysis of chest CT images was performed and compared to healthy controls (**Figure 3**). Among hospitalized and ICU patients, the mean percent of total lung classified as GGO was 13.2% and 28.7%, respectively, and was higher than in ambulatory patients (3.7%, P<0.001 for both comparisons). Although the amount of GGO observed in the ambulatory group was low, it was higher than healthy controls (0.06%; P<0.001). GGO as a percentage of total lung correlated with the percent predicted TLC (ρ = -0.6; P<0.001) and DLCO (ρ = -0.47; P<0.001). DPM analysis revealed that a substantial percentage of total lung was affected by air trapping in all severity groups, and there were no differences in the amount of air trapping across PASC groups. The mean percentage of total lung affected by air trapping was 25.4%, 34.5% and 27.2% in the ambulatory, hospitalized and ICU groups and was higher in ambulatory patients than in healthy controls (7.3%; P<0.001). These differences persisted after adjusting for age and BMI (**Figure S3**).

**Table 2:** Chest computed tomography findings.

| **Table 2: Chest computed tomography findings** |  |  |  |  |
| --- | --- | --- | --- | --- |
| **Finding - no. (%)** | **All Patients**<br>**(N=91)** | **Ambulatory**<br>**(N=59)** | **Hospitalized**<br>**(N=16)** | **ICU**<br>**(N=16)** |
| Air trapping | 50/91 (58%) | 32/56 (57%) | 11/15 (73%) | 7/15 (47%) |
| Ground glass opacities | 46/91 (51%) | 21/59 (36%) | 10/16 (63%) | 15/16 (94%)*** |
| Pulmonary nodule | 32/91 (35%) | 19/59 (32%) | 9/16 (56%) | 4/16 (25%) |
| Nodule type - no. (%) |  |  |  |  |
| Solid | 22/32 (69%) | 13/19 (68%) | 7/9 (78%) | 2/4 (50%) |
| Ground glass | 6/32 (19%) | 5/19 (26%) | 0/9 (0%) | 1/4 (25%) |
| Mixed | 4/32 (12%) | 1/19 (5%) | 2/9 (22%) | 1/4 (25%) |
| Bronchiectasis | 23/91 (25%) | 5/59 (8%) | 7/16 (44%)** | 11/16 (69%)*** |
| Architectural distortion,<br>honeycombing or scar | 22/91 (24%) | 2/59 (3%) | 7/16 (44%)*** | 13/16 (81%)*** |
| Bronchial wall thickening | 7/91 (7%) | 4/59 (7%) | 2/16 (12%) | 1/16 (6%) |
| Lymphadenopathy | 3/91 (3%) | 0/59(0%) | 0/16 (0%) | 3/16 (19%)** |
| Emphysema | 3/91 (3%) | 0/59(0%) | 1/16 (6%) | 2/16 (12%) |
| Interstitial abnormalities | 3/91 (3%) | 0/59(0%) | 2/16 (12%) | 1/16 (6%) |
| Consolidation | 2/91 (2%) | 0/59 (0%) | 0/16 (0%) | 2/16 (12%) |
| Pleural effusion | 0/91(0%) | 0/59(0%) | 0/16 (0%) | 0/16 (0%) |

Imaging data is not available for 8 ambulatory patients and 1 hospitalized patient.
\*\*P<0.01 and \*\*\*P<0.001 compared to ambulatory patients.

**Figure 3:**
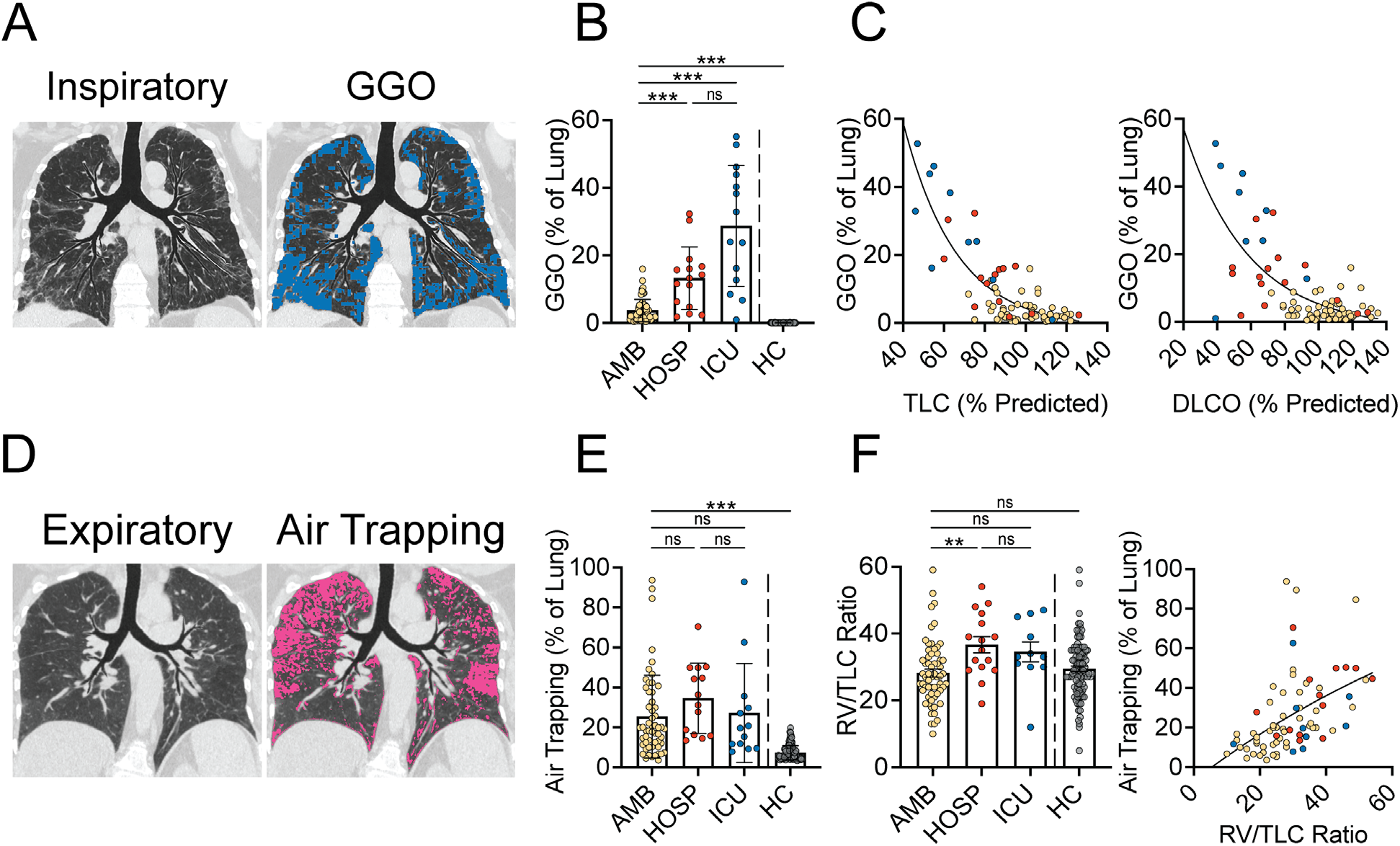
Quantitative chest computed tomography and correlation with pulmonary function. (Panel A) Representative inspiratory chest computed tomography (CT) images (left) obtained at total lung capacity (TLC) and texture analysis map (right) highlighting ground glass opacities (GGO; in blue). (Panel B) Quantification of GGO measured via texture analysis. (Panel C) Correlation of GGO with percent predicted TLC (left) and DLCO (right). (Panel D) Representative expiratory CT images (left) obtained at residual volume (RV) and disease probability measure (DPM) map (right) highlighting air trapping (in pink). (Panel E) Quantification of air trapping measured via DPM. (Panel F) Ratio of RV to TLC by group and correlation of air trapping with RV/TLC. Images (Panels A and D) were prepared using topographic multi-planar reformat (tMPR) rendering, which serves to display the airways and associated parenchyma on the same plane^12,34^. Data are displayed as mean and SEM (Panels B, E and F). Yellow circles, ambulatory; red circles, hospitalized; blue circles, ICU. *Abbreviations*: AMB, ambulatory; HOSP, hospitalized; ICU, intensive care unit; HC, healthy controls. *P<0.05, **P<0.01 and ***P<0.001.

Air trapping can also be detected using lung volume measurements, specifically the RV to TLC ratio (RV/TLC). We therefore assessed RV/TLC using both plethysmography and quantitative CT. Quantitative CT measurement of RV and TLC were obtained during the same effort from which air trapping was measured. The RV/TLC ratio was higher in the hospitalized group compared to the ambulatory group (P<0.01) by plethysmography (**Figure 3**) but was not different across groups when measured by quantitative CT (**Figure S4**). RV/TLC calculated using plethysmography correlated with the presence of air trapping measured by DPM (ρ = 0.6; P<0.001) as did RV/TLC calculated using quantitative CT (ρ = 0.84; P<0.001).

Finally, we assessed whether there was a relationship between the severity of pulmonary function or CT abnormalities with time since acute infection or with respiratory symptoms. No significant correlations were found between spirometry, lung volumes, GGO or air trapping and days since diagnosis (**Figure S5**). PASC patients were separated by quartile using quantitative measures of GGO and air trapping, and the proportion of patients with dyspnea or cough in each quartile was assessed. There were no differences in the proportion of patients with respiratory symptoms across quartiles (**Figure S6**).

## Discussion

This prospective case series describes 100 patients with PASC. Patients were stratified by the highest level of care required during acute illness, and symptoms, pulmonary function testing and chest imaging were performed. As recently reported by others, persistent respiratory symptoms including dyspnea and cough were among the most common symptoms reported by PASC patients in our cohort. Hospitalized and ICU patients were more dyspneic than ambulatory patients as measured by the mMRC score, but the frequency of dyspnea and cough was not different across severity groups. Additionally, PASC patients who required hospitalization or ICU care during acute infection had restrictive physiology and impaired gas exchange on pulmonary function testing, characterized by a reduction in FVC, FEV1, TLC and DLCO. These patients were also more likely to have bronchiectasis and architectural distortion, honeycombing or scar and bronchiectasis on chest CT imaging compared to the ambulatory group. These findings are consistent with the sequelae of severe COVID-19 reported by others^2,3,11,22^.

In contrast to most published reports of post-acute COVID-19, the majority of PASC patients in our cohort had mild disease during acute infection and did not require hospitalization. Spirometry and lung volumes were normal in these patients and were not different than a cohort of healthy control subjects. We observed a higher DLCO in the ambulatory group compared to healthy controls. Increases in DLCO can be driven by increases in pulmonary capillary blood volume, as occurs in asthma and obesity^23^. Approximately a quarter of patients in the ambulatory group had asthma, and the BMI of the ambulatory group was significantly higher than healthy controls. Alternatively, increases in pulmonary capillary blood volume could be driven by endothelial dysfunction, as has been reported following SARS-CoV-2 infection^24,25^.

Analysis of inspiratory and expiratory chest CT images, both by an experienced radiologist and using quantitative image analysis, revealed the presence of GGO and air trapping in ambulatory patients. Compared to hospitalized and critically ill patients, fewer patients in the ambulatory group had GGO on qualitative analysis of chest CT images and a smaller percentage of total lung was classified as GGO by texture analysis. Importantly, texture analysis revealed that GGO as a percentage of total lung was significantly higher in the ambulatory group compared to healthy controls, suggesting there is ongoing lung inflammation, edema or fibrosis in PASC patients. A striking proportion of patients in all three PASC groups had air trapping on qualitative CT analysis. The percent of total lung classified as air trapped using DPM analysis was not different across PASC groups, and there was significantly more air trapping measured by DPM in the ambulatory group compared to healthy controls. Air trapping occurs as a result of partial or complete airway obstruction in regions of the lung. We did not observe airflow obstruction by spirometry in any group, suggesting that air trapping in our cohort is due to involvement of small rather than large airways. Small airways, defined as non-cartilaginous airways with an internal diameter < 2mm, contribute little to total airway resistance and thus, small airway disease is typically not detected by spirometry until a large percentage (>75%) of all small airways are obstructed^26–28^. While small airways are not readily identified by lung imaging, a number studies have established the presence of air trapping on chest CT as a biomarker of functional small airways disease (fSAD)^19–21^. Taken together, our findings suggest that SARS-CoV-2 infection itself leads to fSAD and air trapping, while restrictive lung disease and impairment in gas exchange results from lung injury and ARDS, regardless of the underlying cause.

The angiotensin-converting enzyme 2 (ACE2) receptor, which facilitates SARS-CoV-2 infection, is expressed throughout the airway tract, including in the small airways^29,30^. Thus, the fSAD observed in PASC patients (PASC-fSAD) could result from direct infection of the small airways by SARS-CoV-2, even in patients with mild acute infection. In this case, PASC-fSAD may result from an ongoing injury-repair process, cellular debris and/or abnormal mucus production. Conversely, the immune response induced by SARS-CoV-2 could induce PASC-fSAD even in the absence of direct infection. Regardless of the underlying mechanism, the presence of fSAD months after the acute infection raises concern for airway remodeling and fibrosis. Of note, we did not observe a relationship between pulmonary function tests or quantitative CT measures of GGO or air trapping and time from diagnosis. The median time from diagnosis to pulmonary function measurements and chest CT imaging was approximately 75 days in our cohort. Persistent respiratory abnormalities in this timeframe could represent a prolonged resolution-repair process following SARS-CoV-2 infection and might therefore represent post-infectious bronchiolitis as has been described following other severe viral infections^31,32^. However, small airways disease is also observed as a result of chronic inflammatory disorders such as connective tissue disease and inflammatory bowel disease, immunodeficiency, and following exposure to certain noxious stimuli^26^. In many of these cases, irreversible and sometimes progressive lung impairment is observed. Given the prevalence of PASC among COVID-19 survivors and the proportion of patients that appear to be affected by PASC-fSAD, longitudinal studies aimed at understanding the natural history of disease should be pursued.

We did not observe a correlation between dyspnea and the severity of either GGO or air trapping measured by quantitative CT analysis. This is perhaps not surprising given the relatively small number of patients in our cohort and might suggest that in addition to fSAD, other pathological processes contribute to the respiratory symptoms frequently observed in PASC. Extra-pulmonary involvement, including increased cardiometabolic demand, exertional tachycardia, chronic fatigue and anxiety, have been reported in patients with PASC^33^. Further studies will be needed to more fully understand the mechanisms contributing to dyspnea in post-COVID-19 patients.

### Limitations

This is single center study, and thus the generalizability of our findings may be limited. Furthermore, we only evaluated symptomatic patients, and therefore cannot comment on the prevalence of lung function and imaging abnormalities among all COVID-19 survivors.

Longitudinal assessment will be required to determine whether PASC-fSAD improves over time, or whether it leads to persist or progressive lung disease.

### Conclusions

Our study demonstrates that patients with PASC have a high prevalence of long-lasting air trapping by imaging, regardless of the initial severity of infection. Air trapping is often missed with spirometry but can be detected using inspiratory and expiratory CT imaging and plethysmography. Studies aimed at determining the natural history of PASC-fSAD and the biological mechanisms that underlie these findings are urgently needed in order to identify therapeutic and preventative interventions.

## Data Availability

The individual participant data that underlie the results reported in this Article, as well as the statistical analysis plan, will be made available after deidentification. Researchers who provide a methodologically sound proposal for any purpose may direct proposals to. To gain access, data requestors will need to sign a data access agreement that requires approval by University of Iowa Office of Technology Transfer.

## Acknowledgements

The authors would like to acknowledge the patients who participated in this study. We also thank Kimberly Sprenger, Sue Ellen Salisbury, Maria Aguilar Pescozo, Deborah O’Connell-Moore, and Jarron Atha for their assistance with this study. The authors would also like to acknowledge the contributions of the University of Iowa Institute of Clinical and Translational Science which is supported in part by the National Institutes of Health (UL1TR002537). The views expressed in this article are those of the authors and do not necessarily reflect the position or policy of the Department of Veterans Affairs or the United States Government.

