## Supplementary material for "Small Airways Disease is a Post-Acute Sequelae of SARS-CoV-2 Infection"

### Supplementary Appendix

**Materials and Methods**

**Figure S1.** Multivariate adjusted analysis of pulmonary function testing

**Figure S2.** Representative chest CT images

**Figure S3.** Multivariate adjusted analysis of quantitative chest CT data

**Figure S4.** Correlation between RV/TLC and air trapping

**Figure S5.** Correlation of pulmonary function tests and quantitative chest CT measures with days since diagnosis

**Figure S6.** Respiratory symptoms by severity of quantitative chest CT abnormalities

**Table S1.** Demographic characteristics of healthy subjects

**Table S2.** Clinical characteristics of acute COVID-19

**Table S3.** Clinical characteristics of post-acute COVID-19

**Table S4.** Outcomes

**Supplementary References**

### Materials and Methods

#### Study Design, Definitions and Assessments

During the clinic visit, patients were asked to recount their symptoms during the acute phase of SARS-CoV-2 infection (within 21 days of diagnosis) and whether these symptoms persisted. Symptoms included in the questionnaire were fever (>38°C or 100.4°F), cough, dyspnea, anosmia, dysgeusia, myalgia, upper respiratory tract symptoms, fatigue, nausea, headache, diarrhea, lightheadedness and rash.

The modified Medical Research Council (mMRC) scale scores dyspnea as follows: 0 – dyspnea only with strenuous exercise, 1 – dyspnea when hurrying or walking up a slight hill, 2 – walks slower than people of the same age because of dyspnea or has to stop for breath when walking at own pace, 3 – stops for breath after walking 100 yards or after a few minutes, 4 – too dyspneic to leave house or breathless when dressing[1,2].

Pulmonary function testing, including pre- and post-bronchodilator spirometry and measurement of plethysmographic lung volumes and diffusion capacity, was performed according to American Thoracic Society / European Respiratory Society (ATS/ERS) guidelines[3]. Percent predicted values were determined by comparing measured values to a standard reference cohort (Global Lung Initiative-2012), endorsed by ATS/ERS[4].

Asymptomatic healthy controls (N=106) were enrolled between 2018 and 2019 as part of a separate protocol approved by the institutional review board of the University of Iowa. Healthy control subjects were 20 to 80 years of age, had no prior history of cardiopulmonary disease and were non-smokers, defined as less than 1 pack of cigarettes in their lifetime. All subjects provided written informed consent.

#### Chest Computed Tomography (CT) Acquisition and Analysis

For all healthy control subjects and eighty-seven PASC patients, chest CT scans were obtained on a Siemens SOMATOM Force. The remaining 4 PASC patients were imaged utilizing a Siemens Definition AS+. All scans included iterative reconstruction and dose modulation. Adaptive Multiple Feature Method (AMFM)-based texture analysis was used to quantify ground glass opacities (GGO) and ground glass reticular (GGR) as previously described and validated[5–10]. AMFM analysis was completed in combination with supervised machine learning. For training, CT image textures were selected via a Bayesian Classifier utilizing radiologist defined parenchymal labels in an independent image data set. For the purposes of patient evaluation, GGO and GGR regions were expressed as percent of lung volume at TLC, and the percentages were combined and referred to as GGO. Disease probability measure (DPM) was calculated by Lung Print software (VIDA Diagnostics, Inc, Coralville, Iowa) to quantify air trapping[11]. The probability of air trapping is estimated through warping of the inspiratory and expiratory image and quantifying the voxel-to-voxel difference in Hounsfield Units (HU) between the two CT volume images such that the probability is inversely proportional to the relative differences in HU. If a region is labeled as emphysema on inspiration, it is eliminated from being labeled as air trapped on expiration. Air trapping measured by DPM has also been referred to as functional small airways disease (fSAD)[11–13].

**Figure S1**

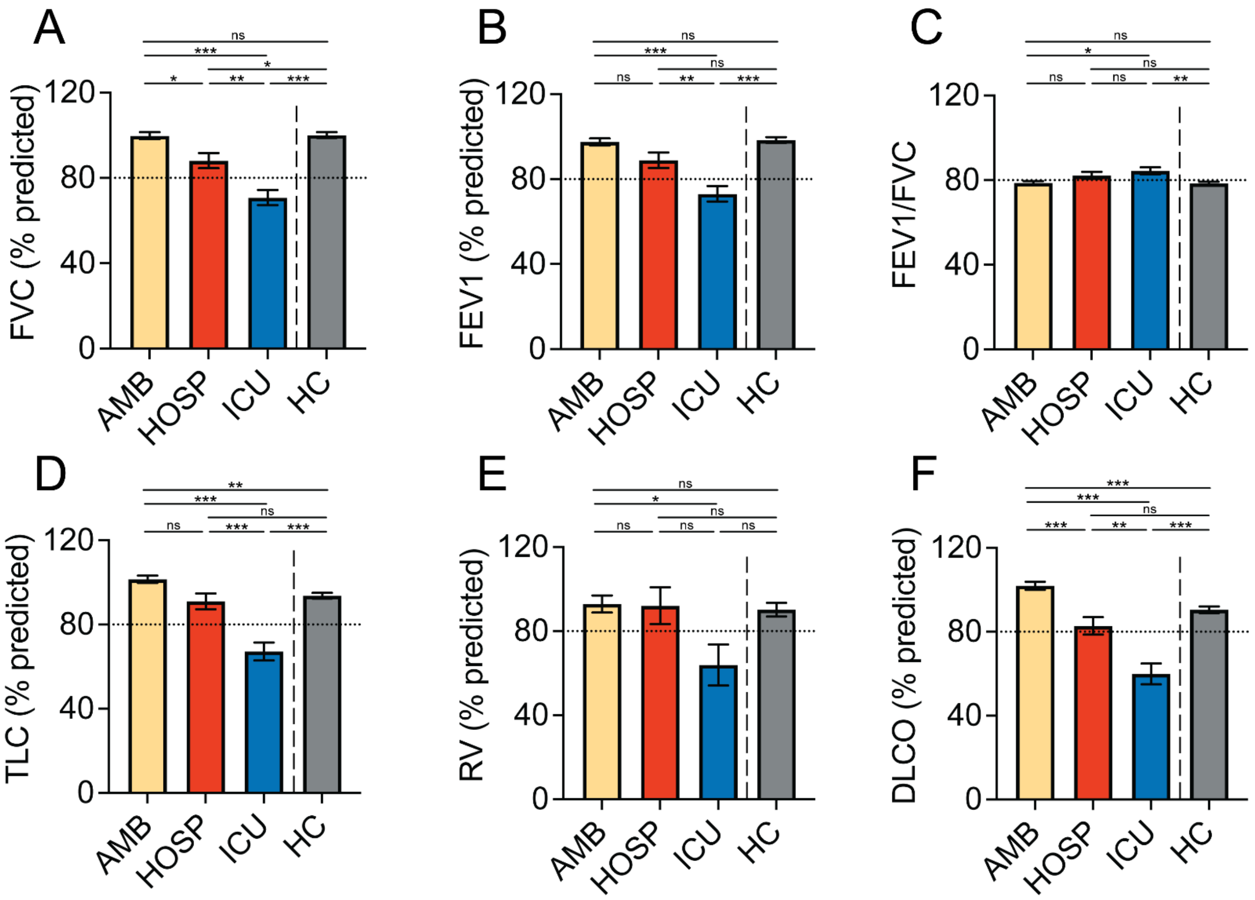

**Figure S1: Multivariate adjusted analysis of pulmonary function testing**

Shown are the pulmonary function testing data by group, adjusted for age and body mass index (BMI) using multivariate linear regression. (Panel A) Percent predicted forced vital capacity (FVC). (Panel B) Percent predicted forced expiratory volume in 1 second (FEV1). (Panel C ) FEV1/FVC. (Panel D) Percent predicted total lung capacity (TLC). (Panel E) Percent predicted residual volume (RV). (Panel F) Percent predicted diffusion capacity for carbon monoxide. Data are displayed as least square mean (LSmean) with standard error measurement (SEM). Horizontal dashed lines indicate the lower limit of normal. *P<0.05, **P<0.01 and ***P<0.001.

*Abbreviations*: AMB, ambulatory; HOSP, hospitalized; ICU, intensive care unit; HC, healthy controls

**Figure S2**

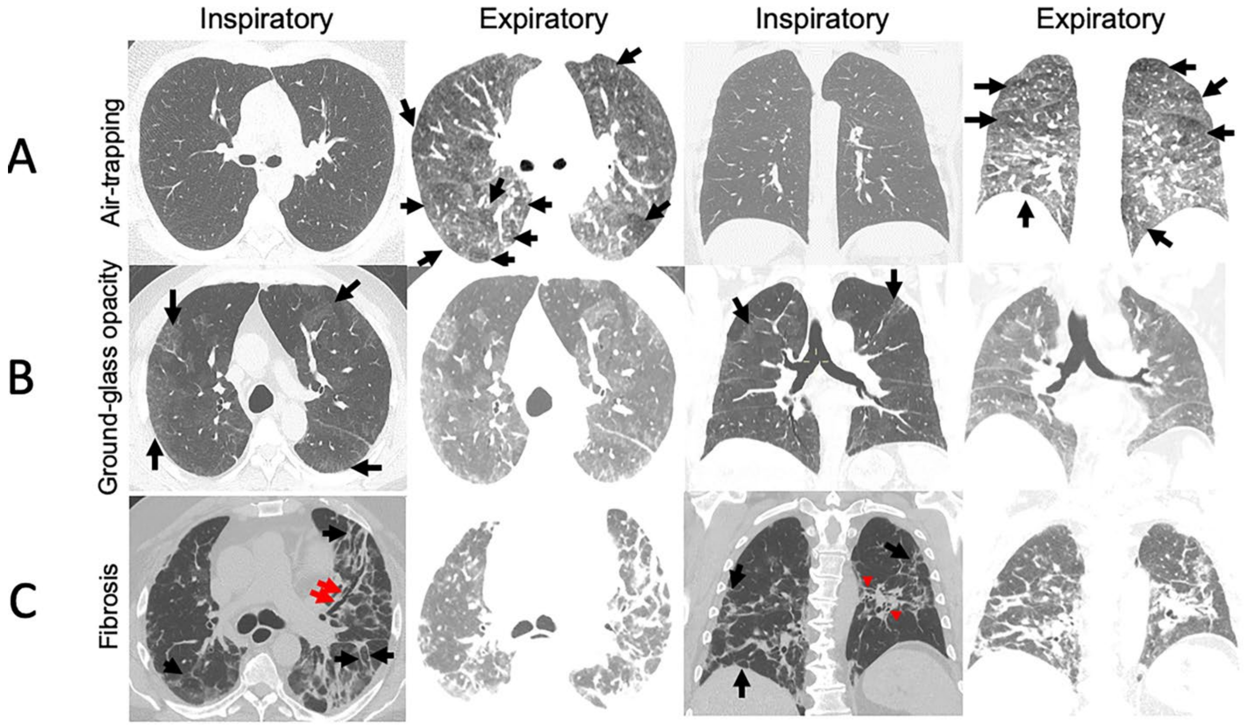

**Figure S2: Representative chest CT images**

Shown are representative chest CT images from separate patients following COVID-19 infection. Each row represents images from a single patient and includes axial and coronal slices obtained during inspiration to total lung capacity (TLC) and expiration to residual volume (RV). (Panel A) Inspiratory and expiratory images obtained seven months following COVID-19 diagnosis. Inspiratory images are normal but expiratory images reveal multiple areas of air trapping (black arrows). (Panel B) Extensive bilateral ground glass opacities (GGO) are demonstrated on the inspiratory images (black arrows) obtained two months after the diagnosis of COVID-19. These areas have higher attenuation on expiratory images, confirming the presence of GGO. (Panel C) Images obtained five weeks after diagnosis of COVID-19 demonstrate fibrosis with linear areas of interstitial thickening (black arrows), bronchiectasis (red arrows) and volume loss with crowding of the broncho-vascular structures (red arrowheads).

**Figure S3**

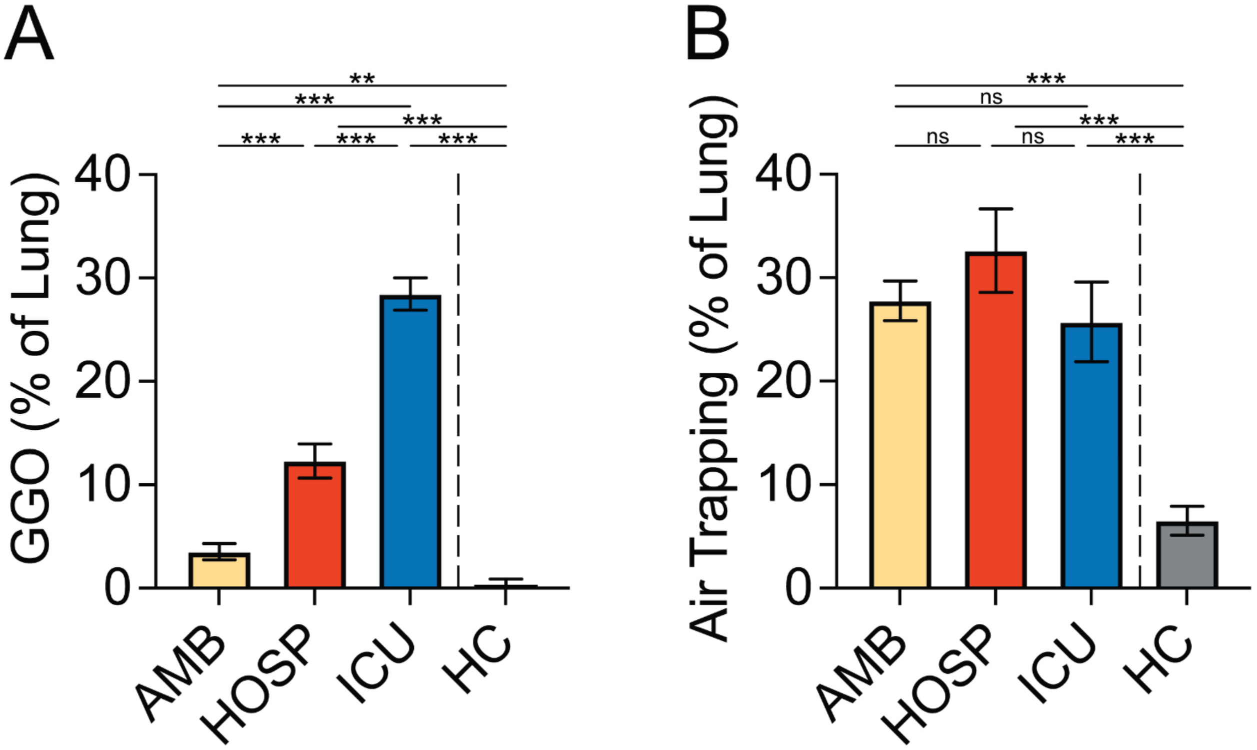

**Figure S3: Multivariate adjusted analysis of quantitative chest CT data**

Shown are the quantitative chest CT data by group, adjusted for age and BMI using multivariate linear regression. (Panel A) Quantification of GGO measured via texture analysis. (Panel B) Quantification of air trapping measured by DPM. Data are displayed as LSmean and SEM. *Abbreviations*: AMB, ambulatory; HOSP, hospitalized; ICU, intensive care unit; HC, healthy controls. **P<0.01 and ***P<0.001.

**Figure S4**

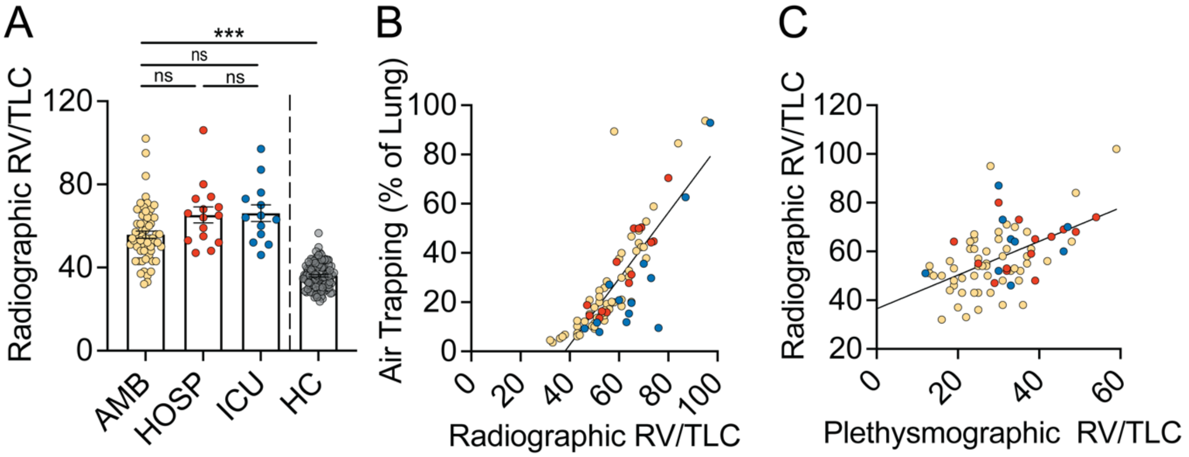

**Figure S4: Correlation between RV/TLC and air trapping**

(Panel A) RV/TLC measured using quantitative chest CT. (Panel B) Correlation between radiographic RV/TLC and air trapping. (Panel C) Correlation between radiographic and plethysmographic RV/TLC. Yellow circles, ambulatory; red circles, hospitalized; blue circles, ICU. *Abbreviations*: RV, residual volume; TLC, total lung capacity. ***P<0.001.

### Figure S5

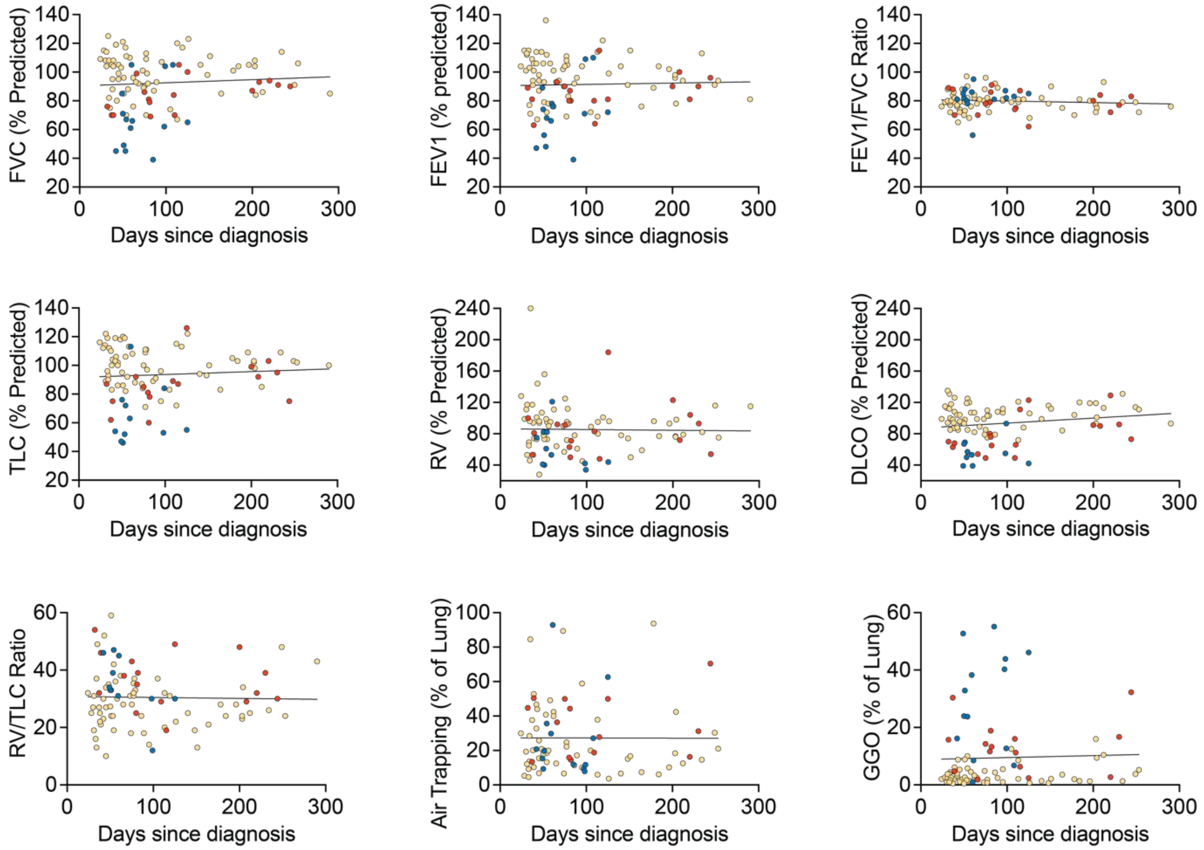

**Figure S5: Correlation of pulmonary function tests and quantitative chest CT measures with days since diagnosis**

Shown are the correlations between pulmonary function tests (FVC, FEV1, FEV1/FVC, TLC, RV, DLCO, RV/TLC), GGO and air trapping with days since diagnosis. Yellow circles, ambulatory; red circles, hospitalized; blue circles, ICU. *Abbreviations*: FVC, forced vital capacity; FEV1, forced expiratory volume in 1 second; RV, residual volume; TLC, total lung capacity; DLCO, diffusion capacity for carbon monoxide.

**Figure S6**

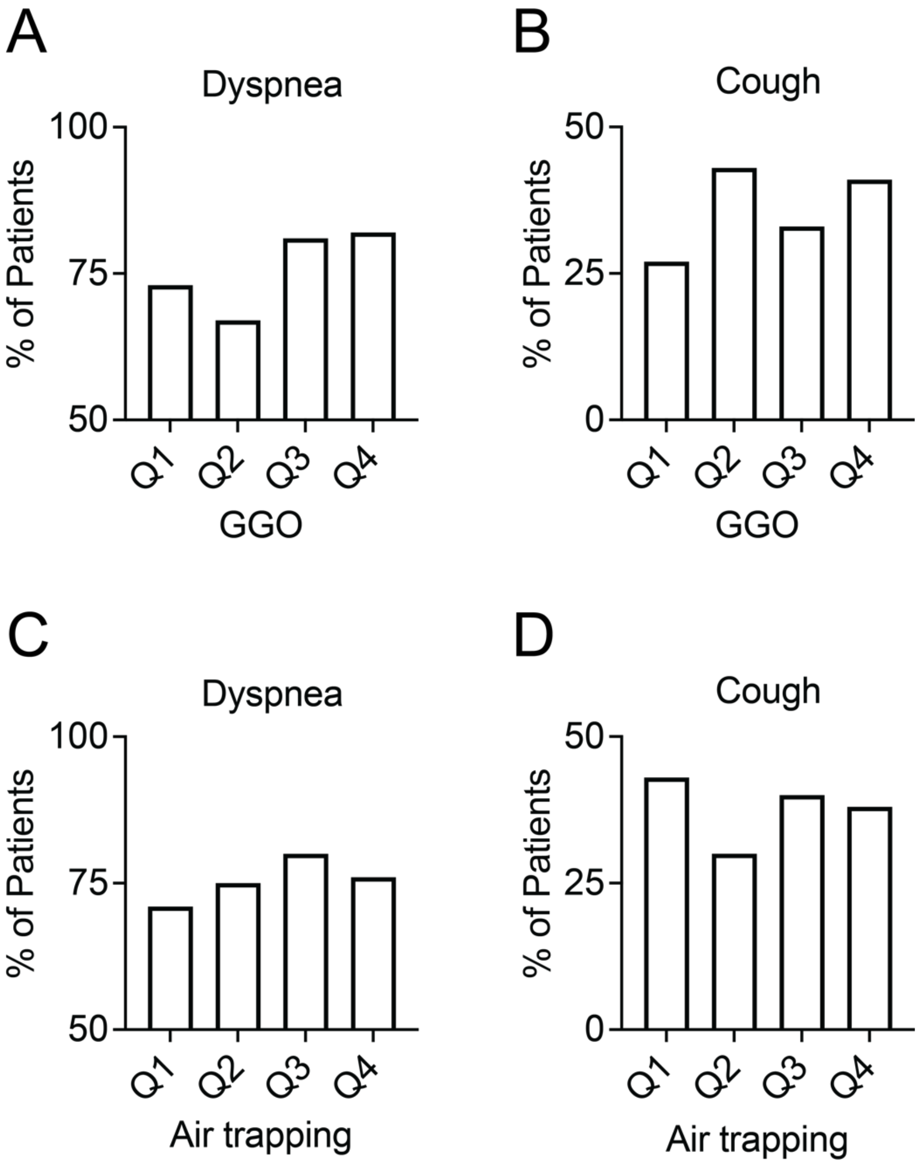

**Figure S6: Respiratory symptoms by severity of quantitative chest CT abnormalities**

(Panel A) Percentage of patients with dyspnea by quartile of GGO as a percentage of total lung. (Panel B) Percentage of patients with cough by quartile of GGO as percentage of total lung. (Panel C) Percentage of patients with dyspnea by quartile of air trapping percentage of total lung. (Panel D) Percentage of patients with cough by quartile of air trapping percentage of total lung.

**Table S1: Demographic characteristics of healthy subjects**

| **Table S1: Demographic characteristic of healthy subjects** | | |
|---|---|---|
| **Characteristic** | **Healthy Subjects (N=106)** | **Post-COVID-19 Patients (N=100)** |
| Age, median (IQR) - yr | 47.5 (30.8-57.3) | 48 (36.3-60.5) |
| Sex - no. (%) | | |
| Male | 46 (43.4%) | 34 (34%) |
| Female | 60 (56.6%) | 66 (66%) |
| Race and ethnic group - no. (%)[#] | | |
| White, non-Hispanic | 97 (91.5%) | 85 (85%) |
| Black, non-Hispanic | 3 (2.8%) | 5 (5%) |
| Hispanic or Latino | 6 (5.7%) | 10 (10%) |
| Body-mass index, median (IQR)[§] | 25.4 (22.7-28.1)*** | 31.5 (26.6-38.1) |

*Abbreviations*: IQR, interquartile range

[#]Race and ethnic group were reported by the patient.

[§]The body-mass index (BMI) is the weight in kilograms divided by the square of the height in meters.

***P<0.001 compared to post-COVID-19 patients.

**Table S2: Clinical characteristics of acute COVID-19**

| **Table S2: Clinical characteristics of acute COVID-19** | | | | |
|---|---|---|---|---|
| **Characteristic** | **All Patients (N=100)** | **Ambulatory (N=67)** | **Hospitalized (N=17)** | **ICU (N=16)** |
| Symptoms - no. (%) | | | | |
| Fatigue | 82/98 (83.7%) | 58/67 (86.6%) | 10/16 (62.5%) | 14/15 (93.3%) |
| Dyspnea | 82/99 (82.3%) | 53/67 (79.1%) | 15/17 (88.2%) | 14/15 (87.5%) |
| Cough | 70/98 (71.4%) | 48/67 (71.6%) | 11/16 (68.8%) | 11/15 (73.3%) |
| Myalgias | 61/98 (62.2%) | 45/67 (67.2%) | 6/16 (37.5%) | 10/15 (66.7%) |
| Upper respiratory symptoms | 59/98 (60.2%) | 47/67 (70.1%) | 7/16 (43.8%) | 5/15 (33.3%)* |
| Fever | 57/98 (58.2%) | 37/67 (55.2%) | 10/16 (62.5%) | 10/15 (66.7%) |
| Anosmia | 48/98 (49%) | 39/67 (58.2%) | 2/16 (12.5%)** | 7/15 (46.7%) |
| Dysgeusia | 44/98(44.8%) | 34/67 (50.7%) | 3/16 (19.8%)* | 7/15 (46.7%) |
| Diarrhea | 40/98 (40.8%) | 28/67 (41.8%) | 7/16 (43.8%) | 5/15 (33.3%) |
| Lightheadedness | 31/98 (31.6%) | 22/67 (32.8%) | 6/16 (37.5%) | 3/15 (20%) |
| Rash | 3/98 (3.1%) | 2/67(3.0%) | 1/16 (6.7%) | 0/15(0%) |
| Laboratory Data[#] | | | | |

| | | | | |
|---|---|---|---|---|
| Highest white-cell count, median (IQR) - per mm3 | -- | -- | 6940 (4250-9650) | 16100 (14550-26975)††† |
| Neutrophils - % | -- | -- | 69.9% (64.7-81) | 76.9% (70.8-84.9) |
| Neutrophils, median (IQR) - per mm3 | -- | -- | 2600 (1600-6380) | 11750 (10282-16917)††† |
| Lymphocytes - % | -- | -- | 15.1% (7.9-27) | 9.1% (6.6-16.2) |
| Lymphocytes, median (IQR) - per mm3 | -- | -- | 710 (600-950) | 1513 (1040-2200)† |
| Neutrophil to lymphocyte ratio | -- | -- | 4.33 (2.29-10.08) | 9.18 (4.58-15.86) |
| Lowest white-cell count, median (IQR) - per mm3 | -- | -- | 3480 (2250-5725) | 7000 (5850-10175)††† |
| Neutrophils - % | -- | -- | 60.1% (49.9-74.8) | 75.1% (62.9-83.2) |
| Neutrophils, median (IQR) - per mm3 | -- | -- | 2690 (1000-3730) | 5010 (4785-7770)††† |
| Lymphocytes - % | -- | -- | 25.5% (15.8-3.9) | 14.2% (10.1-22.1) |
| Lymphocytes, median (IQR) - per mm3 | -- | -- | 800 (510-1830) | 1040 (587.5-1300) |
| Neutrophil to lymphocyte ratio | -- | -- | 2.44 (1.25-4.64) | 6.59 (4.37-10.24)† |

| | | | | |
|---|---|---|---|---|
| Lowest platelet count, median (IQR) - per mm3 | -- | -- | 172,000 (137,500-266,500) | 189,000 (105,000-266,500) |
| Lowest hemoglobin, median (IQR) - g/dl | -- | -- | 11.6 (10.5-12.8) | 9.8 (7.3-12.5) |
| Highest blood urea nitrogen, median (IQR) - mg/dl | -- | -- | 18 (14.5-25.3) | 39.5 (28.3-46.8)††† |
| Highest serum creatinine, median (IQR) - mg/dl | -- | -- | 1.0 (0.8-1.2) | 1.1 (0.9-1.6) |
| Highest aspartate aminotransferase, median (IQR) - U/liter | -- | -- | 35 (27-63) | 48.5 (34.8-63) |
| Highest alanine aminotransferase, median (IQR) - U/liter | -- | -- | 29 (25-90) | 45.5 (27-63.3) |
| Highest bilirubin, median (IQR) - mg/dl | -- | -- | 0.4 (0.3-0.7) | 0.6 (0.4-1.1) |
| Highest D-dimer, median (IQR) - mg/ml | -- | -- | 0.58 (0.42-1.36) | 6.66 (2.19-26.75)†† |
| Highest fibrinogen - median (IQR) - mg/dl | -- | -- | 516 (447-669) | 665.5 (594.8-843.8)†† |
| Duration of admission - days, median (IQR) | | | | |
| Hospital | -- | -- | 4.0 (2.5-7) | 18.5 (10.8-42.3)††† |

| | | | | |
|---|---|---|---|---|
| ICU | -- | -- | -- | 13.5 (6.5-22.8) |
| Need for supplemental oxygen - no. (%)§ | 26 (26%) | 0 (0%) | 10 (58.8%) | 16 (100%) |
| COVID-19 treatments - no. (%) | | | | |
| Corticosteroids | 36 (36%) | 11 (16.4%) | 11 (64.7%)*** | 14 (87.5%)*** |
| Antibiotics | 18 (18%) | 6 (9.0%) | 7 (41.2%)** | 5 (31.3%) |
| Remdesivir | 16 (16%) | 0 (0%) | 5 (29.4%)*** | 11 (68.8%)*** |
| Convalescent plasma | 13 (13%) | 1 (1.5%) | 3 (17.6%)* | 9 (56.3%)*** |
| Other experimental medication | 6 (6%) | 0 | 3 (17.6%)** | 3 (18.8%)** |
| ICU Level Therapies - no. (%) | | | | |
| High-flow oxygen | -- | -- | -- | 14 (87.5%) |
| Invasive mechanical ventilation | -- | -- | -- | 11 (68.8%) |
| Prone position | -- | -- | -- | 9 (56.3%) |
| Vasopressors | -- | -- | -- | 9 (56.3%) |
| Neuromuscular blockade | -- | -- | -- | 5 (31.2%) |
| CPAP or noninvasive positive pressure ventilation | -- | -- | -- | 4 (25%) |
| Extracorporeal membrane oxygenation | -- | -- | -- | 3 (18.8%) |
| COVID-19 complications - no. (%) | | | | |

| | | | | |
|---|---|---|---|---|
| Tracheostomy | 3 (3%) | 0 (0%) | 0 (0%) | 3 (18.8%)** |
| Need for renal replacement | 2 (2.0%) | 0 (0%) | 0 (0%) | 2 (12.5%)** |
| Pneumothorax or pneumomediastinum | 1 (1.0%) | 0 (0%) | 0 (0%) | 1 (6.3%) |
| Thromboembolic disease | 5 (5%) | 2 (3.0%) | 2 (11.8%) | 1 (6.3%) |

*Abbreviations:* CPAP, continuous positive airway pressure; IQR, interquartile range

#Laboratory data is missing for 5 hospitalized patients and is incomplete for 5 hospitalized patients and 3 ICU patients.

§Median maximal supplemental oxygen for hospitalized patients was 2 liters per minute (IQR, 0-2). Median maximal fraction of inspired oxygen for ICU patients was 1.0 (IQR, 0.65-1.0). Supplemental oxygen dose was missing for 3 hospitalized patients.

*p < 0.05, **p < 0.01, and ***p < 0.001 compared to ambulatory patients.

†p < 0.05, ††p < 0.01, and †††p < 0.001 compared hospitalized patients.

**Table S3: Clinical characteristics of post-acute COVID-19**

| Table S3: Clinical Characteristics of Post-Acute COVID-19 | | | | |
|---|---|---|---|---|
| **Characteristic** | **All Patients (N=100)** | **Ambulatory (N=67)** | **Hospitalized (N=17)** | **ICU (N=16)** |
| Symptoms - no. (%) | | | | |
| Dyspnea | 73/100 (73%) | 45/67 (67.2%) | 14/17 (82.4%) | 14/16 (87.5%) |
| Fatigue | 56/100 (56%) | 39/67 (58.2%) | 10/17 (58.8%) | 7/16 (43.8%) |
| Cough | 34/100 (34%) | 21/67 (31.3%) | 4/17(23.5%) | 9/16 (56.3%) |
| Myalgias | 27/100 (27%) | 19/67 (28.4%) | 4/17 (23.5%) | 4/16 (25%) |
| Upper respiratory symptoms | 21/100 (21%) | 14/67(20.9%) | 4/17 (23.5%) | 3/16 (18.8%) |
| Lightheadedness | 19/100 (19%) | 13/67 (19.4%) | 4/17 (23.5%) | 2/16 (12.5%) |
| Anosmia | 17/100 (17%) | 14/67 (21%) | 0/17 (0%) | 3/16 (18.8%) |
| Dysgeusia | 14/100 (14%) | 10/67 (14.9%) | 1/17 (5.9%) | 3/16 (18.8%) |
| Diarrhea | 10/100 (10%) | 8/67 (11.9%) | 1/17 (5.9%) | 1/16 (6.3%) |
| Subjective fever | 5/100 (5%) | 3/67(4.5%) | 1/17 (5.9%) | 1/16 (6.3%) |
| Rash | 3/100 (3.0%) | 3/67 (4.5%) | 0/17(0%) | 0/16 (0%) |
| Vital signs, median (IQR)[#] | | | | |
| Temperature - °C | 36.5 (36.2-36.8) | 36.5 (36.3-36.8) | 36.5 (36.2-37.1) | 36.4 (36.2-36.8) |
| Pulse – beats per minute | 81 (71-93) | 80 (71-92) | 81 (66-92) | 90 (70-100) |
| Systolic blood pressure - mmHg | 130 (120-139) | 128 (118-138) | 134 (122-140) | 136 (125-145) |
| Diastolic blood pressure - mmHg | 75 (69-84) | 76 (69-83) | 70 (67-79) | 81 (71-88) |
| Respiratory rate – breaths per minute | 16 (16-18) | 16 (16-18) | 16 (16-18) | 16 (16-19) |

| | | | | |
|---|---|---|---|---|
| SpO2 -% | 98 (96-99) | 98 (97-99) | 98 (96-98) | 98 (96-99) |
| Supplemental oxygen | | | | |
| Ongoing use - no. (%) | 11 (11%) | 0 (0%) | 2 (11.8%) | 9 (56.3%)*** |
| Rate - median (IQR), liters per minute | 0 (0-0) | 0 (0-0) | 0 (0-0) | 1 (0-2) |
| Laboratory Data§ | | | | |
| White blood cells, median (IQR) - per mm3 | 7400 (6100-9100) | 7300 (6200-8800) | 6500 (5550-9100) | 9100 (7900-11500) |
| Neutrophils, median (IQR) - % | 63.5% (56.8-68) | 63% (52.5-67) | 64% (59-71) | 69.5% (58.5-75.8) |
| Neutrophils, median (IQR) - per mm3 | 4425 (3278-6168) | 4370 (3270-5995) | 4010 (3335-5075) | 6070 (3953-8825) |
| Lymphocytes, median (IQR) - % | 25% (21.8-31) | 26% (23.5-34) | 24% (16.5-27.5) | 19.5% (17-29.3) |
| Lymphocytes, median (IQR) - per mm3 | 1870 (1458-2490) | 1970 (1540-2530) | 1440 (1170-2080) | 1895 (1560-2585) |
| Neutrophil to lymphocyte ratio | 2.49 (1.81-3.13) | 2.45 (1.52-2.85) | 2.40 (2.04-4.79) | 3.29 (1.68-4.35) |
| Platelet count, median (IQR) - per mm3 | 270,000 (243,000-325,000) | 273,000 (243,500-322,500) | 266,000 (175,500-365,000) | 266,000 (246,500-361,500) |
| Hemoglobin, median (IQR) - g/dl | 13.6 (12.9-15) | 13.8 (13.3-14.8) | 13.2 (11.7-15.4) | 13.2 (11-14.9) |
| Blood urea nitrogen, median (IQR) - mg/dl | 13 (10-17) | 13 (10-17) | 14 (12-19) | 12 (8-18) |
| Serum creatinine, median (IQR) - mg/dl | 0.8 (0.7-0.9) | 0.8 (0.8-0.9) | 0.9 (0.7-1.0) | 0.8 (0.6-1.0) |

| | | | | |
|---|---|---|---|---|
| Aspartate aminotransferase, median (IQR) - U/liter | 22 (18-29.5) | 21 (17-28) | 28 (23.8-37) | 23.5 (16.5-28.8) |
| Alanine aminotransferase, median (IQR) - U/liter | 21 (16-35) | 19 (15.5-34) | 27 (21-43.3) | 24.5 (13.8-37) |
| Bilirubin, median (IQR) - mg/dl | 0.4 (0.3-0.5) | 0.4 (0.3-0.5) | 0.4 (0.3-0.8) | 0.4 (0.4-0.5) |

*Abbreviations*: IQR, interquartile range; SpO2, percent saturation of oxygen in the blood measured by pulse oximetry

#Vital sign data is missing for 2 ambulatory and 3 hospitalized patients and is incomplete for 14 ambulatory, 1 hospitalized and 2 ICU patients.

§Laboratory data from 45 ambulatory patients, 13 hospitalized patients and 10 ICU patients is shown.

***p < 0.001 compared to ambulatory patients.

**Table S4: Outcomes**

| Table S4: Outcomes | | | | | |
|---|---|---|---|---|---|
| **Outcome** | **All Post-COVID-19 (N=100)** | **Ambulatory (N=67)** | **Hospitalized (N=17)** | **ICU (N=16)** | **Healthy Control (N=106)** |
| Score on the mMRC scale, median (IQR)[#] | 2 (0-2) | 1 (0-2) | 3 (1-3)* | 2 (1.3-3) | -- |
| FeNO, median (IQR) - ppb[§] | 14 (11-22.3) | 13 (11-20) | 19 (13.5-46.5) | 16 (8.8-25.5) | -- |
| FVC - % of predicted value | | | | | |
| Pre-bronchodilator, median (IQR)[^] | 95 (81-106) | 103 (88.5-109) | 85 (71.5-93.3)** | 65.5 (48-90)*** | 100 (93.8-108) |
| Post-bronchodilator, median (IQR)[^^] | 97 (86-107) | 104 (90-108) | 88 (75.8-93)** | 66 (52.8-90.5)*** | -- |
| Bronchodilator response, median (IQR) - % | 1 (-2-3) | 0 (-2.5-3) | 1.5 (-1-7.3) | 2 (0.5-9) | -- |
| FEV1 - % of predicted value | | | | | |
| Pre-bronchodilator, median (IQR)[^] | 93 (78-104) | 99 (89.5-106) | 87 (80-91.5)* | 71.5 (54-80.8)*** | 98 (91-107) |

| | | | | | |
|---|---|---|---|---|---|
| Post-bronchodilator, median (IQR)^^ | 96 (85-108) | 102 (92-110) | 86 (82.5-91.5)*** | 73 (53.3-81.3)*** | -- |
| Bronchodilator response, median (IQR) - % | 3 (1-6) | 3 (1-5.5) | 3 (0.8-8) | 5 (2-7) | -- |
| FEV1/FVC | | | | | |
| Pre-bronchodilator, median (IQR)^ | 0.8 (0.76-0.84) | 0.79 (0.76-0.82) | 0.79 (0.73-0.87) | 0.83 (0.81-0.86) | 0.79 (0.75-0.82) |
| Post-bronchodilator, median (IQR)^^ | 0.82 (0.75-0.86) | 0.82 (0.77-0.86) | 0.79 (0.71-0.85) | 0.85 (0.76-0.86) | -- |
| FEF25-75% - % predicted value | | | | | |
| Pre-bronchodilator, median (IQR)^ | 100 (77.5-127) | 91 (67-106.5) | 86 (58.8-105.8) | 105.5 (43.5-118.5) | -- |
| Post-bronchodilator, median (IQR)^^ | 82 (75-86) | 82 (77-86) | 79 (70.8-85.3) | 84.5 (76-86) | -- |
| Bronchodilator response, median (IQR) - % | 12 (0.3-22.8) | 14 (2.5-23.5) | 6.5 (-7.5-14.8) | 10 (-8.5-28.5) | -- |
| TLC, median (IQR) - % of predicted value¶ | 96 (83.5-109) | 102 (90-111) | 87 (76-95)** | 55 (52-76)*** | 95 (88-102) |
| RV, median (IQR) - % of predicted value¶ | 82 (64.3-98.8) | 91 (73-99.5) | 82 (56-98) | 53 (41-82)** | 89 (76.8-106.8) |

| | | | | | |
|---|---|---|---|---|---|
| RV/TLC, median (IQR)¶ | 30 (24-36.8) | 27 (22-33.2) | 37 (29-45)** | 33 (30-47) | 29 (23.8-35) |
| DLCO, median (IQR) - % of predicted value¶¶ | 96 (79-111.5) | 103 (93-113)††† | 73 (64-91.5)*** | 54 (41.3-67.5)*** | 88 (84-97.3) |

*Abbreviations:* DLCO, diffusing capacity for carbon monoxide; FeNO, fraction of exhaled nitric oxide; FEF25-27%, forced expiratory flow at 25-75% of forced vital capacity; FEV1, forced expiratory volume in 1 second; FVC, forced vital capacity; IQR, interquartile range; mMRC, modified Medical Research Council; RV, residual volume; TLC, total lung capacity

#mMRC data is missing for two ambulatory patients.

§FeNO data is shown for 51 ambulatory, 14 hospitalized and 6 ICU patients.

^Pre-bronchodilator data is missing for two ambulatory and two ICU patients.

^^Post-bronchodilator data is missing for 4 ambulatory, 2 hospitalized and 6 ICU patients.

¶Lung volume data is missing for 4 ambulatory, 1 hospitalized and 5 ICU patients.

¶¶DLCO data is missing for 4 ambulatory and 6 ICU patients.

*p < 0.05, **p < 0.01, and ***p < 0.001 compared to ambulatory patients.

†††p < 0.001 compared to healthy control subjects.
